# Comprehensive characterization of *Candida* isolates in a given geographical area for the determination of prevalence and drug sensitivity

**DOI:** 10.1101/2022.02.25.22270246

**Authors:** Tamanna Pamnani, Saifali Agarwal, Jayati Chourasia, Rishika Sinha, Pankaj Choudhary, Ravi Ranjan K Niraj, Ramjas Nagar, Sandeep K Shrivastava, Aakanksha Kalra

## Abstract

**Purpose:** An alarming increase in Candidiasis infections has been observed worldwide and in India. This increase is attributed to increase in immune-compromised individuals and an increase in plethora of species causing the disease. The emergence of drug resistance of isolates has further worsened the situation.

**Materials & Methods:** *Candida* isolates causing infections were obtained from patients and evaluated for their macroscopic and microscopic characteristics by colony morphology and staining procedures. The patients’ demography was also analyzed to identify any correlation between the isolates. Doubling time was determined to analyze growth characteristics of these isolates. Isolates were also characterized biochemically for their ability to assimilate carbon and nitrogen sources and for fermentative capabilities. Additionally, drug sensitivity profiles of these isolates were analyzed towards Azoles.

**Results:** Demographic analysis of the isolates suggested that all age groups were affected by *Candida via* both albicans and non-albican species. The infections were not gender biased but majority isolates were obtained from urine samples suggesting *Candida* as an important species causing urine-genital infections. Macroscopic and microscopic analysis showed cream colored circular colonies with smooth surface and entire margins with cells in single and budding stage. The doubling time ranged between 40 mins to 180 mins with 81 mins being the average. Biochemical characterization showed sucrose to be the most metabolizable sugar with maximum fermentative capacity with glucose. Surprisingly, very low nitrogen assimilation capacity was observed with all the nitrogen sources tested (nitrate, urea and glycine). Drug sensitivity towards Azoles suggested almost 50% and 90% isolates were resistant to Fluconazole and Itraconazole respectively, the most common Azoles used against fungal infections in the current scenario.

**Conclusions:** The results obtained from the study suggested differential characteristics of the isolates towards various parameters thereby indicating the relevance of isolate characterization, for appropriate control and prevention of the disease.

**Graphical abstract:** 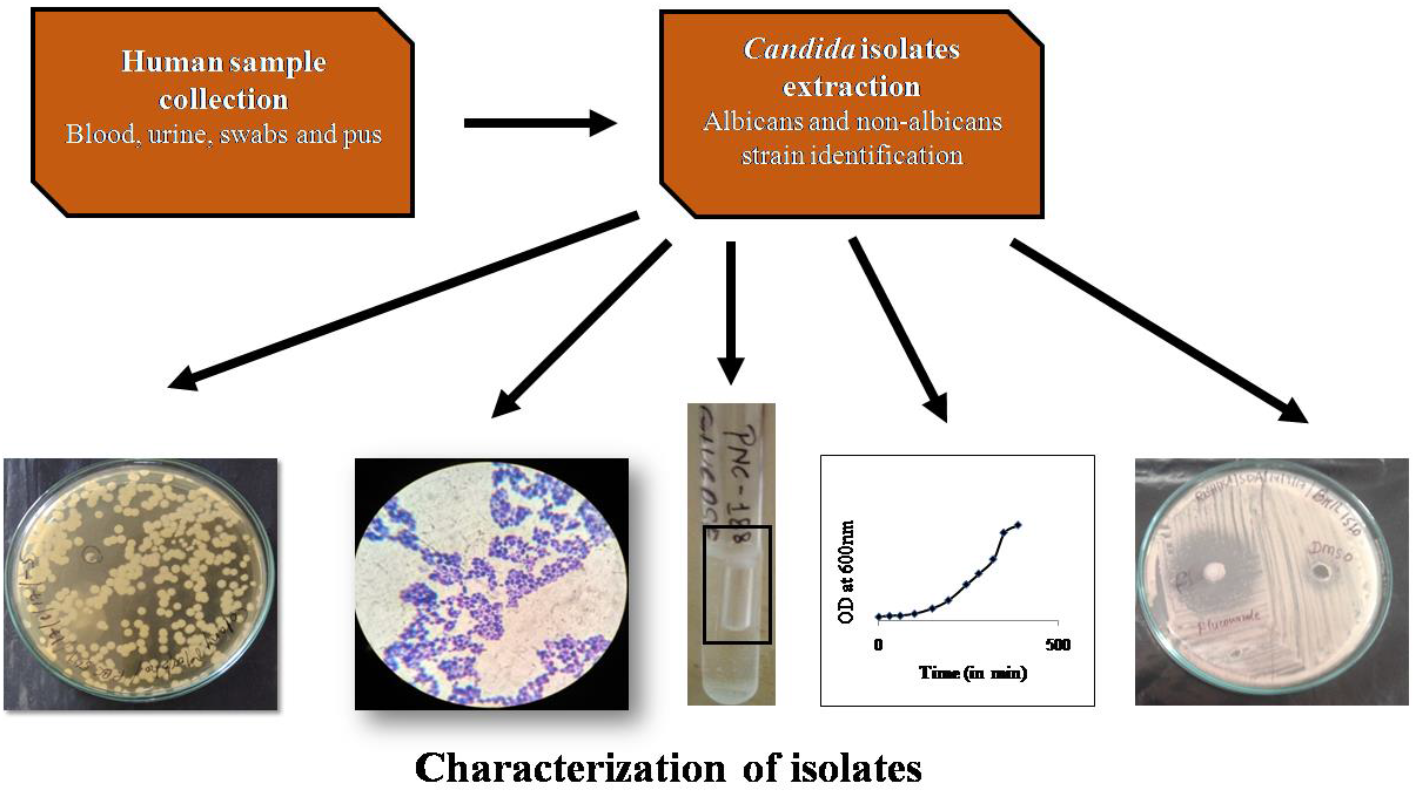

## Introduction

A wide variety of fungi have been responsible for causing life threatening infections worldwide leading to annual deaths. *Candida* is one of the commonest causes of these infections particularly bloodstream infections which have been rising annually [1]. Members of the genus *Candida* use biotic or abiotic surfaces to colonize and spread the disease in hospitals [2]. The National Nosocomial Infections Surveillance System (NNISS) has considered *Candida* species as fourth most common nosocomial bloodstream pathogen [3]. Also, there has been a drastic increase in HCAIs*-* Health care associated infections, at a prevalence rate of 9.1%, for which *albicans* and *glabrata* species are major causative agents in addition to bacterial species [4].

Numerous studies have shown that of the 150 known species of *Candida*, only a handful are known to cause infections ranging from sub-cutaneous to systemic in immuno-compromised individuals. Previously, *C. albicans* was assumed to be responsible for majority of these infections (∼80%) with other non-albicans species such as *glabrata, krusei, tropicalis*, etc. However, infections caused by these species are increasing day by day as these have been isolated from patients and have been marked as pathogens [5, 6]. Also, there has been emergence of novel species such as *C. auris* causing the disease [7-9].

These studies suggest that identification and characterization of strains is of utmost importance for the prevention and control of the disease. In lieu of this, the present study is aimed at recovery of *Candida* species causing infections in Rajasthan. These isolates were then comprehensively characterized as described in the upcoming sections.

## Materials & Methods

### *Candida* isolates and culture conditions

The study comprises of 30 *Candida* isolates obtained from human samples procured for diagnostic purposes at Dr. B. Lal Clinical Laboratory Pvt. Ltd., Jaipur. The isolates were kindly provided by MCRD-CIRD (Microbial Culture Repository Division - Center for Innovation, Research and Development), research division at BCL. Of these, only 23 isolates could be revived on Yeast Extract Peptone Dextrose (YEPD) media (HiMedia Laboratories, India) and were included in the study. All the isolates were maintained on YEPD medium (unless specified) at 30°C. Appropriate approvals have been taken for the same.

### Macroscopic & microscopic characterization

The colony morphology was analyzed by standard spread plate technique. Primary inoculation of isolates was followed by ten-fold serial dilution in saline which were then spread on YEPD agar and incubated at 30°C for 24 hrs. The grown colonies were analyzed for morphological characteristics.

For microscopic analysis, microbes were visualized for shape, size and multiplication state via staining with Indian ink, KOH, lactophenol cotton blue, Gram stain and Giemsa stain.

### Growth characterization

The growth characteristics were determined to analyze their multiplicative abilities by standard procedures. Briefly, primary inoculation was done in YEPD broth and incubated at 30°C for 24 hrs. 1% inoculum from primary culture was used for secondary culture and optical density of this culture was taken at 600nm (0hr absorbance) [10, 11] and then at regular intervals to monitor the growth of isolates. The growth curves were plotted with respect to time to determine the log phase and determine the doubling time using the following equation:

Doubling time = Log_10_2/r, where r = log x/ time in hrs for which x = final OD of log phase/initial OD of log phase

### Biochemical characterization

The carbon and nitrogen assimilation capacity towards respective sources affect infection rate [12] and thus was analyzed as per standard procedures. Briefly, for carbon assimilation, YNB broth (yeast nitrogen base), containing different carbon sources (glucose, lactose, sucrose and ethanol) at 2% concentration was used. YNB broth without any carbon source was used as negative control. The growth of isolates in different sources was analyzed after 24 hrs via optical density at 600nm and was compared to 0hr OD and with negative control. Similarly, for nitrogen assimilation capacity, a nitrogen free media, YCB broth (yeast carbon base) was used and the nitrogen sources analyzed included nitrate, urea and glycine. YCB broth was used as negative control.

Fermentation media (NaCl, peptone and either of the carbon sources) were used for the analysis as per standard procedures. Media was inoculated with the isolate followed by insertion of a sterile Durham tube in inverted position and incubation at 30°C for 24 hrs. The isolate growth was determined by presence of cell pellet at the bottom of the tube and the fermentation capacity was measured by the bubble formed in Durham tube.

### Drug sensitivity profiles towards Azoles

Azoles have been the most commonly used antifungal drugs owing to low cost, limited toxicity and oral administration. Therefore, the drug sensitivity of these isolates was determined towards Fluconazole and Itraconazole, the two most common Azoles, as per standard procedures with NCCLS guidelines by agar well diffusion assay [13]. Drugs were dissolved in dimethyl sulfoxide (DMSO) at a concentration of 12.5mg/ml. The isolates were initially inoculated in Sabouraud Dextrose Broth (SDB) and incubated at 30°C for 24 hrs. These cultures were then diluted to an OD of 0.2 and swabbed on Sabouraud Dextrose Agar (SDA) plates. Wells were punctured in these plates and 50μL of the drug was added. Wells containing DMSO were used as negative control. Plates were incubated for 24 hrs at 30°C and zone of inhibition was measured by zone measuring scale.

## Results & Discussion

### Demographic and morphological analysis

The details of the isolates obtained from MCRD-CIRD, BCL are summarized in Table S1 which includes isolate ID, age, gender of the patient, sample type and classification of the isolate (albican vs non-albican). The demographic analyses are shown in Figure 1a which clearly shows that the disease affects all age groups at a similar rate with the youngest patient of 03 days age while the oldest of 80 years. However, the results suggest that the middle age group (31 to 60 years) is slightly more affected which can be attributed to increased cases of genital candidiasis. This is consistent with the previous studies showing higher infection rates of bacterial vaginosis in middle age groups more specifically reproductive age groups. Though *Candida* is one of the major causes for these infections, studies have failed to show such a correlation (14). Also, it was observed that the infection rates are similar in both males and females (Figure 1a). The ratio of infected males to females is 0.91:1. Multiple studies pertaining to UTI infections in females in post-menopausal stage suggest controversial results but presence of *Candida* was ensured in post-menopausal infections in 2014 thereby further confirming such infections in all age groups (15). The isolates used in the study have been obtained from multiple samples including urine, blood, vaginal swabs or pus. Figure 1(a) clearly shows that majority of the isolates were obtained from urine samples, suggesting *Candida* to be one of the major causes of urinary infections. A study in Yemen has suggested bacterial vaginosis to be the major cause of reproductive infections followed by vulvovaginal candidiasis (16). Though both bacteria and fungi are known to be associated with UTIs, multiple studies have claimed the decrease in bacterial infections with a rise in fungal infections for the same (17). The spread of fungal infections has also been attributed to sexual routes as their presence has been observed in penile tissue (16). Unfortunately, a large proportion of isolates (45%) in our study were obtained from blood samples suggesting severity of the disease. The species identification is done by germ tube assay performed at BCL and the information was kindly shared along with the isolates’ details. A positive germ tube test is an indicative of the albican species recognized as production of germ tube on incubation of the isolates in the presence of human serum at 37°C. The analysis of infectious isolates as albicans or non-albicans showed the infections to be equally caused by both categories (Figure 1a). This is both surprising and concerning since till date albican species was responsible for majority of the infections. A study on hospital acquired infections over a period of 7 years in the US has claimed *Candida* particularly albican species to be a major cause of these infections (18). Though non-albican species have been emerging as a cause of UTIs, *Candida albicans* still remains the leading cause for the fungal UTIs (19). However, the results in this study state otherwise. Recent studies have suggested the increase in infections caused by non-albican species such as *glabrata, tropicalis*, etc. [6, 20]. Increase in emergence of infections caused by *C. auris* is another such example [7]. Geographical variations have also emphasized the distribution of disease-causing species with *C. albicans* as predominantly associated in North and Central Europe and the USA while non-albicans species predominate in Asia, South Europe, and South America (21).

**Figure 1:**
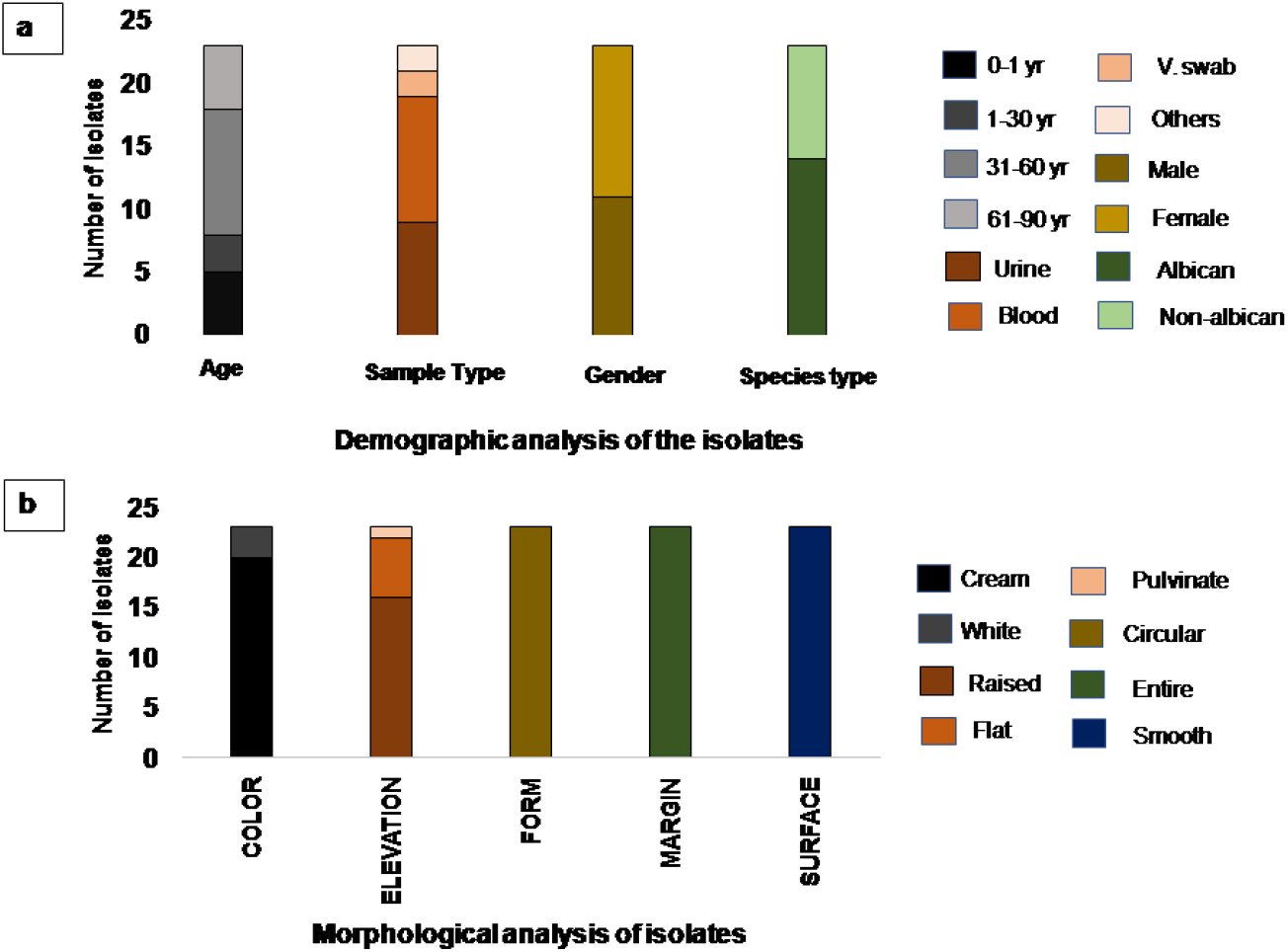
**<u>(a) Demographic analysis of isolates:</u>** The proportion of isolates is categorized on the basis of age groups, sex-ration, sample type and albican-nonalbicans species. **(b) <u>Macroscopic characterization:</u>** The colony morphology of all isolates compiles color, form, margin, surface and elevation of the isolates.

The morphological characterization of the colonies on YEPD agar was analyzed in terms of color, form, margin, surface and elevation. Representative images of a few isolates for colony morphology are shown in Figure S1a. Similar colony morphologies were observed for almost all the isolates with minor variations, characteristics for the same are compiled in Table S2. All the colonies were cream in color, typical of yeast colonies with two isolates showing white circular colonies with smooth surface and entire margins (Figure 1b).

The microscopic analysis was done for the analysis of size and shape of the isolates via five different staining procedures (Indian ink, KOH staining, LPCB staining, Gram stain and Giemsa stain). As observed in all the staining procedures, the cells were oval in shape and were either in single celled form or in the budding stage (Figure S1b).

### Growth characterization

Growth of the isolates was analyzed spectrophotometrically by measuring the absorbance of the culture medium at 600nm with respect to time. Doubling time of the isolates was calculated by analysis of the log phase of each isolate using the equation stated in materials and methods. It was observed that doubling time of these isolates varied from 40 mins to 180 mins with the average doubling time to be 81 mins (Table S3). On the basis of doubling time shown in Table 1, these isolates were categorized in four classes as fast growing, average, slow growing and very slow growing isolates. It was observed that the majority of the isolates belonged to either fast growing (approximately 60%) or average growing category (approximately 30%) suggesting the severity of infection dissemination. Only 1 isolate in each category was observed with slow and very slow growing doubling time. The growth curves of these isolates represent the standard microbial growth curve consisting of lag, log, stationary and decline phase with variations in log phase which are the cause of differential doubling times.

**Table 1:** Growth characterization of isolates: All isolates are grouped in four categories as Fast growing, average growing, slow growing and very slow growing.

| **SR.NO.** | **DOUBLING TIME RANGE** | **ISOLATE NO.** |
| --- | --- | --- |
| 1. | **Fast growing**<br>**(41-80 mins)** | **14** |
| 2. | **Average growing**<br>**(81-120 mins)** | **7** |
| 3. | **Slow growing**<br>**(121-160 mins)** | **1** |
| 4. | **Very Slow growing**<br>**(161-200 mins)** | **1** |

### Biochemical characterization

The isolates were analyzed for their carbon and nitrogen assimilation capacities and fermentative capacities. Four different carbon sources including monosaccharide (glucose), disaccharide (lactose), polysaccharide (sucrose) and an alcohol (ethanol) were used for the study. Results showed differential carbon assimilation capacities as the fold change in growth of isolates in the presence of sucrose is very high as compared to other carbon sources (Table S4). In addition to sucrose, fold change in growth is higher with glucose as compared to lactose or ethanol. The average fold change in growth of the isolates was found to be 3.27 fold for sucrose and 2.47 fold, 1.41 fold and 1.44 fold for glucose, lactose and ethanol respectively (Figure 2a).

**Figure 2:**
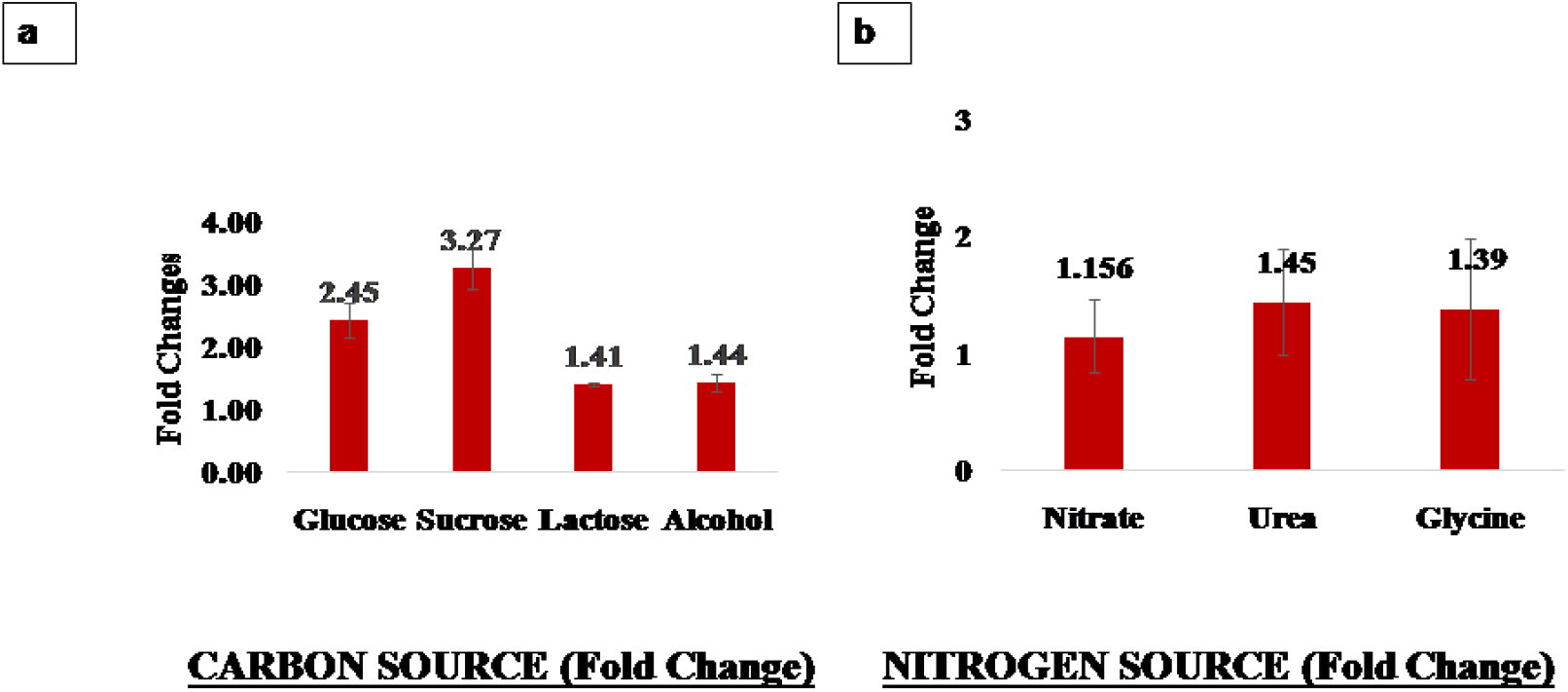
**<u>(a) Average fold change (carbon):</u>** The figure represent the average fold change of four carbon sources (glucose, lactose, sucrose & alcohol) by isolates. **<u>(b) Average fold change (Nitrogen):</u>** The figure represent the average fold change of three nitrogen sources (nitrate, urea & glycine) by isolates.

Similarly, nitrogen assimilation capacity was evaluated with urea, nitrate and glycine (Table S5). When the growth of isolates in nitrogen free media was compared to the growth in presence of nitrate, urea and glycine, an average fold change of -1.15,-1.45 and -1.39 was observed for these sources respectively (Figure 2b). Results showed that the nitrogen assimilation capacity of isolates was very less compared to the carbon assimilation capacities with very low capacity even with simplest amino acid, glycine. Analysis of assimilation capacity of *S. cerevisiae* isolates (a close relative of *Candida*) for nitrogen sources suggested that neither urea nor amino acids alone can be the sole nitrogen source (22). Urea metabolism via Dur1, 2 is important for resistance to innate host immunity in *C. albicans* infections (23).

Another remarkable characteristic of yeast is their fermentation capacity which was evaluated for these isolates as per standard protocols described in the methods section (Figure S2). The bubble length for the isolates towards different carbon sources is compiled in Table 2 and results clearly show that almost all the isolates could ferment glucose. In accordance with the carbon assimilation results, fermentation capacity was observed with sucrose as against lactose and ethanol. It was also observed that the fermentation capacity was highest for NHC-143 which showed highest carbon assimilation capacity for both glucose and sucrose.

**Table 2:** Fermentation capacity of isolates: The table shows the length of air bubble formed during the fermentation of four different carbon sources (glucose, lactose, sucrose & alcohol) by isolates.

| SR.NO. | ISOLATE ID | CARBON SOURCE |  |  |  |
| --- | --- | --- | --- | --- | --- |
|  |  | GLUCOSE | LACTOSE | SUCROSE | ALCOHOL |
| 1 | VDN-804 | 2 cm | 0 | 0 | 0 |
| 2 | BHIL-1385 | 2.3 cm | 0 | 1.7 cm | 0 |
| 3 | NHC-143 | >2.5 cm | 0 | 1.5 cm | 0 |
| 4 | SHC-13 | 1.4 cm | 0 | 0.5 cm | 0 |
| 5 | BHIL-1408 | 2.3 cm | 0 | 2.4 cm | 0 |
| 6 | BHIL-1147 | 2.3 cm | 0 | 1.4 cm | 0 |
| 7 | BHIL-1081 | >2.5 cm | 0 | 1.6 cm | 0 |
| 8 | KNC-81 | 0.9 cm | 0 | 1.2 cm | 0 |
| 9 | VDN-1785 | 1.4 cm | 0 | 0.9 cm | 0 |
| 10 | RPC-662 | 0 | 0 | 0.3 cm | 0 |
| 11 | NHC-193 | 1.5 cm | 0.3 cm | 1.9 cm | 0 |
| 12 | BHIL-1510 | 0.5 cm | 0 | 0 | 0 |
| 13 | SNC-16 | 2 cm | 0.2 cm | 0 | 0 |
| 14 | CDC-382 | 0.4 cm | 0 | 0 | 0 |
| 15 | PNC-188 | 1 cm | 0 | 0.3 cm | 0 |
| 16 | CO-283 | 0.6 cm | 0 | 0 | 0 |
| 17 | PBC-569 | 0 | 0 | 0 | 0 |
| 18 | CO-217 | >2.5 cm | 0 | >2.5 cm | 0 |
| 19 | CO-279 | 0.4 cm | 0 | 0 | 0 |
| 20 | SMS-60 | >2.5 cm | 0 | 0 | 0 |
| 21 | PBC-383 | >2.5 cm | 0 | 0 | 0 |
| 22 | BNC-707 | 0.9 cm | 0 | 0.2 cm | 0 |
| 23 | RPC-450 | >2.5 cm | 0 | 1.3 cm | 0 |

### Drug sensitivity profiles

We tested the sensitivity of *Candida* isolates towards Fluconazole and Itraconazole (most common Azoles administered towards *Candida* infections) by well diffusion assay (Figure S3). It was observed that almost 50% samples showed sensitivity towards Fluconazole while only 2 samples were sensitive to Itraconazole stating 90% resistance (Table S6). Further analysis of the sensitivity pattern of isolates obtained from different type of samples showed no significant difference in the sensitivity pattern (Figure 3). The high resistance of the isolates towards Azoles can be attributed to the long-term exposure of the drug. It has also been observed that the emergence of resistance towards Azoles is infection dependent, with candidemia patients showing the least resistance while oropharyngeal candidiasis showing the highest resistance (20). On the contrary, studies show that fluconazole is still administered alone or in combinations against *Candida* infections particularly UTIs owing to its pharmacokinetics since it is observed to be concentrated maximally in the urine (19, 24). It is both surprising and disappointing since numerous studies have shown that itraconazole is more effective than fluconazole for long term infections. The discrepancy observed in this study could be attributed to the low sample size. Multiple studies in the country and elsewhere have shown emergence of drug resistance towards Azoles [20]. Analysis of the strains and isolates in a respective area are of utmost importance to contain the spreading infections by opportunistic pathogens.

**Figure 3:**
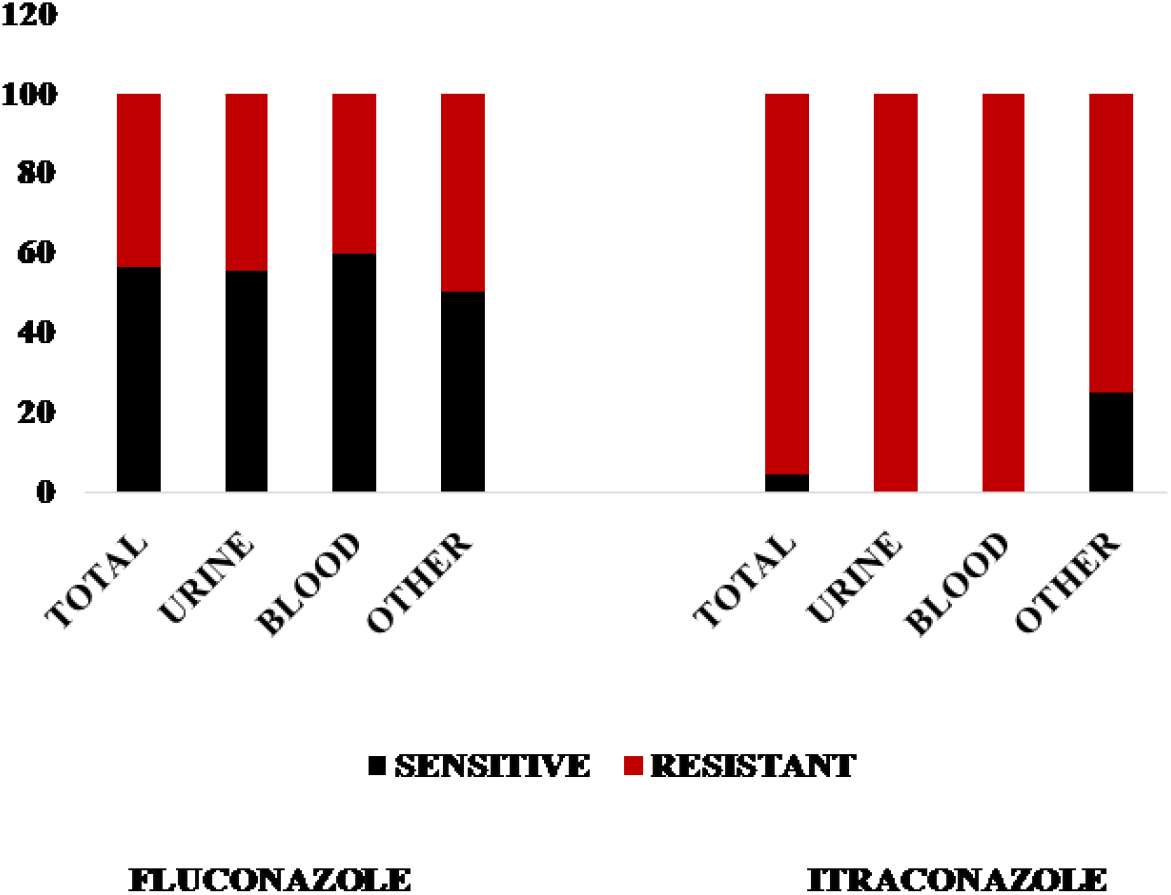
Drug sensitivity analysis: The figure shows drug sensitivity profiles of isolates towards the two Azoles: Fluconazole and Itraconazole.

## Conclusion

A total of 30 isolates were provided for the study of which only 23 could be revived. The demographic analysis of these isolates suggested the spread of infections to all age groups irrespective of gender. The increased proportion of infections by non-albican species is alarming which is consistent with earlier studies [25, 26]. The isolates showed differential growth patterns with doubling time ranging from 40 mins to 180 mins. Surprisingly, sucrose was the most metabolizable sugar promoting the isolate growth with glucose being the highest fermentable sugar. None of the nitrogen sources analyzed could be efficiently metabolized by isolates. Majority of the isolates were resistant to both the Azole drugs analyzed. This is quite disturbing since inspite of resistance observed towards these drugs worldwide, Fluconazole & Itraconazole remain the most commonly administered drugs. The results have shown that characterization of isolates causing infections is of utmost importance for prevention & control of the disease particularly in the pandemic era where the use of steroids has increased extensively promoting opportunistic infections.

## Data Availability

All data produced in the present work are contained in the manuscript

## Declarations

### Consent to participate

The authors have complete consent for the study. **Consent for publication:** The authors have complete consent for publication. **Availability of data and material:** Not Applicable

### Authors’ Contribution

TP performed the experiments and wrote the manuscript; SA, JC, RS, PC performed the experiments; RRKN performed data analysis and manuscript editing; RN, SKS provided the isolates; AK designed the research, planned the experiments & wrote the manuscript.

## Acknowledgments

The authors acknowledge Intramural Research Scientific Committee, Dr. B. Lal Institute of Biotechnology, Jaipur for funding the research.

